# Cross-border transmissions of delta substrain AY.29 during Olympic and Paralympic Games

**DOI:** 10.1101/2021.10.31.21265711

**Authors:** Takahiko Koyama, Reitaro Tokumasu, Kotoe Katayama, Ayumu Saito, Michiharu Kudo, Seiya Imoto

## Abstract

Tokyo Olympic and Paralympic Games, postponed for COVID-19 pandemic, were finally held in summer of 2021. Just before the games, alpha variant was being replaced with more contagious delta variant (B.1.617.2). AY.4 substrain AY.29, which harbors two additional characteristic mutations of 5239C>T (NSP3 Y840Y) and 5514T>C (NSP3 V932A), emerged in Japan and became the dominant strain in Tokyo by the time of the Olympic Games. As of October 18, 98 AY.29 samples are identified in 16 countries outside of Japan. Phylogenetic analysis and ancestral searches identified 46 distinct introductions of AY.29 strains into those 16 countries. United States has 44 samples with 10 distinct introductions, and United Kingdom has 13 distinct AY.29 strains introduced in 16 samples. Other countries or regions with multiple introductions of AY.29 are Canada, Germany, South Korea, and Hong Kong while Italy, France, Spain, Sweden, Belgium, Peru, Australia, New Zealand, and Indonesia have only one distinct strain introduced. There exists no unambiguous evidence that Olympic and Paralympic Games induced cross-border transmission of the delta substrain AY.29. Since most of unvaccinated countries are also under sampled for genome analysis with longer lead time for data sharing, it will take longer to capture the whole picture of cross-border transmissions of AY.29.

## Introduction

Long awaited Tokyo 2020 Olympic and Paralympic Games were postponed for a year due to COVID-19 pandemic. Despite the overwhelming opposing Japanese public opinions, parties including International Olympic Committee (IOC), International Paralympic Committee (IPC), Japanese Government, and Tokyo Metropolitan Government decided to hold the events in summer 2021, starting July 23 and August 24, respectively without spectators in the venues. They made efforts to reduce number of visitors outside of Japan to minimize the risk of importing exogenous novel SARS-CoV-2 strains; as a result, it was substantially reduced to 54,250 from the pre-pandemic estimate of 180,000(1, 2).

Just before the Olympic Games began, in Japan, alpha variant (PANGO lineage(3): B.1.1.7) was being replaced by delta variant, which harbors T478K and L452R mutations in spike protein, with surge of patients due to highly infectious nature of the delta variant(4) (Figure 1A). Japanese Government declared the state of emergency in many prefectures including Tokyo and neighboring prefectures to mitigate the risk of potential healthcare system collapse. Combination of reduction of mobilities, mask compliance, and increase in vaccination rate appeared to have attributed to the significant reduction of positive cases by the end of September as shown in Figure 1B. In fact, reproduction number had already fallen below 1 in Tokyo by mid-August(5), just one week after the end of the Olympic Games.

**Figure 1:**
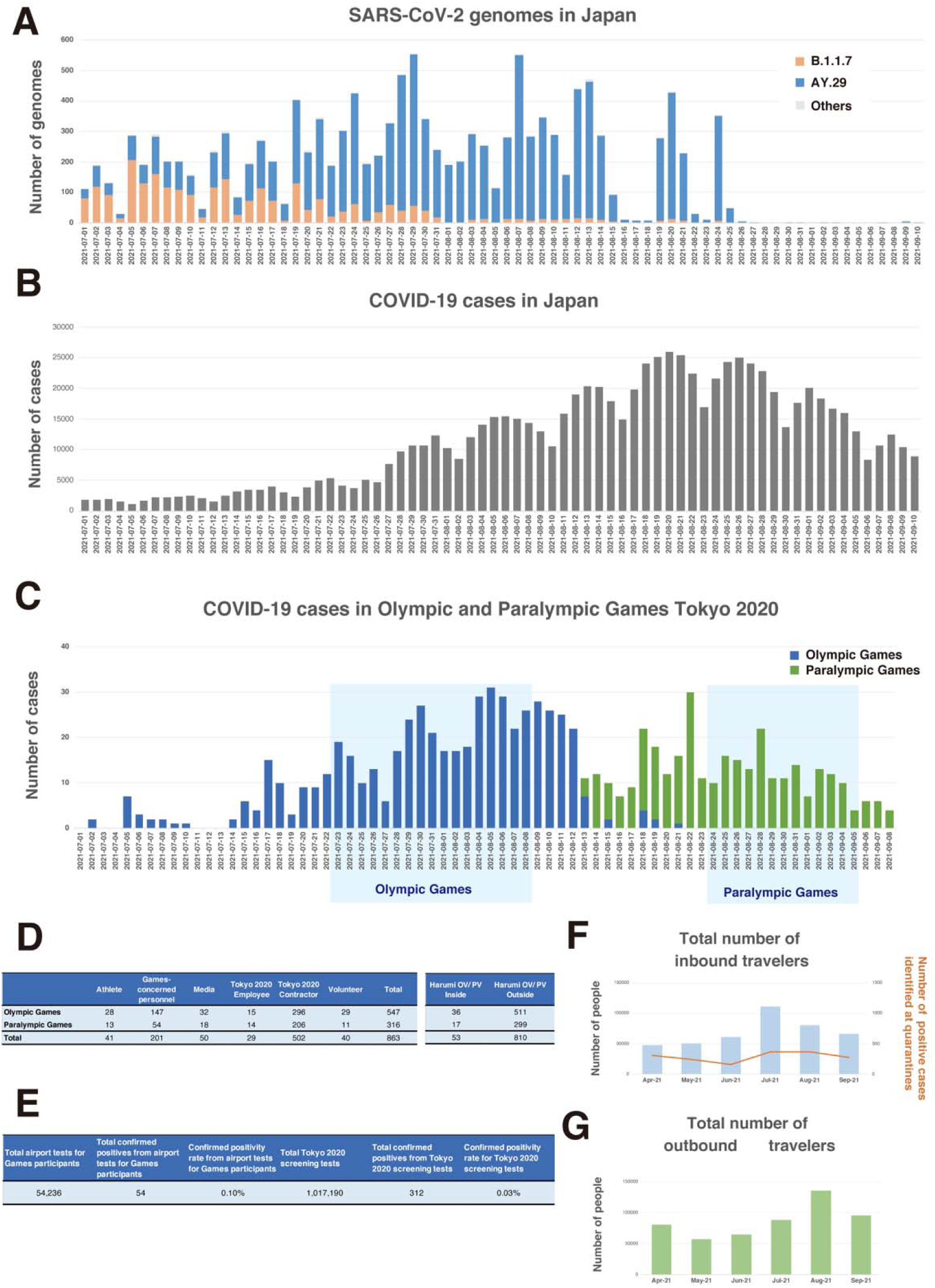
A. Histogram of strains by lineages in Tokyo. B. Number of daily new COVID-19 positive cases in Japan C. Number of daily new COVID-19 positive cases among Olympic and Paralympic participants. D. Statistics of COVID-19 positive cases in the participants E. Statistics of COVID-19 tests performed for the participants. F. Number of inbound travelers to Japan between April and September 2021. G. Number of outbound travelers from Japan between April and September 2021.

Despite the 72-hour testing requirement, Japanese airport quarantine stations identified 55 positive cases at the border control among overseas participants. During the period, total of 863 positive cases were identified(6) (Figure 1C, D). Out of 863 positive cases, 174 cases belong to the Olympic overseas visitors and 80 cases belong to Paralympic overseas visitors. Therefore, majority of the positive cases belong to Japanese residents, such as contract workers and volunteers.

There have been two major concerns for the events regarding COVID-19 since athletes from 205 countries or regions compete in the events. First, Japanese citizens were afraid of novel exogenous strains will be introduced into Japanese population by the participants from abroad, who are waived for any self-quarantine(7). Secondly, variants of concerns (VoC) and variants of interest (VoI) strains besides novel strains, are exported back with the returning participants to the unvaccinated regions. Vaccination rates of low-income countries are still below 5%(8) and introduction to highly infective strains can make devastating outcomes in these areas.

In this study, we have analyzed the SARS-CoV-2 strains transmitted to outside of Japan during the time of the Olympic and Paralympic Games.

## Methods

To perform our analysis, 3,920,975 full genomes extracted from human subjects, were downloaded from GISIAD(9, 10) and National Center for Biotechnology Information (NCBI) up to October 6, 2021. 2,598,663 met a data quality criterion of less than 200 bp gap in an entire genome. Furthermore, latest AY.29 genomes were downloaded from GISAID on October 19, 2021. However, there are some AY.29 strain genomes are misclassified to AY.4 and some AY.4 strains were misclassified to AY.29. To rectify misclassification issues, 5514T>C (NSP3 V932A) and synonymous mutation of 5239C>T were used as criteria and definition of AY.29 for the further analysis.

Variant annotation of SARS-CoV-2 genomes was performed as described in our previous report(11). In a nutshell, the strain was first aligned in a pairwise manner with NC_045112 SARS-CoV-2 reference genome using Needleman-Wunsh algorithm(12) using EMBOSS needle with a gap penalty of 100 and extension penalty of 0.5. From the pairwise alignment, differences with the reference genome were extracted as genome changes and subsequently, annotated for the types of mutation and amino acid changes if any.

Phylogenetic analysis was carried out on all the AY.29 strains exported and randomly selected 1,000 AY.29 genomes from Japan. This selection consists of genomes with full collection date information and no unknown bases or gaps. We used Bayesian evolutionary analysis by sampling trees (BEAST), version 2.6.6. We first aligned sequences using MAFFT(13) and employed Hasegawa-Kishino-Yano mutation model(14), with the strict clock mode. For an overseas sample without exact date information, day 15 was assigned. For visualization, we used ggtree(15) with country and subtype information.

Similarly, ancestral strains search was performed for the overseas AY.29 samples using the maximum variant approach(16). An ancestral strain should have a subset of mutations of the child strain. Among such ancestral strain, one with the maximum common variants is considered as an immediate ancestor or a parent in an ideal situation. Nevertheless, in many samples in delta variants, spike G142D, T95I, and 156_158delinsG are missing because of sequencing artifacts(17). To find proper ancestral strains, these mutations were overlooked for our ancestral searches in this study.

## Results

It is apparent that delta substrain AY.29, which harbors two characteristic mutations of 5239C>T (NSP3 Y840Y) and 5514T>C (NSP3 V932A)(18), was the dominant strain during the time of Olympic and Paralympic Games in Tokyo as shown in Figure 2. Figure 1A shows that alpha strain (B.1.1.7) was being replaced by AY.29 around the end of June and early July before the Game started on July 23.

**Figure 2:**
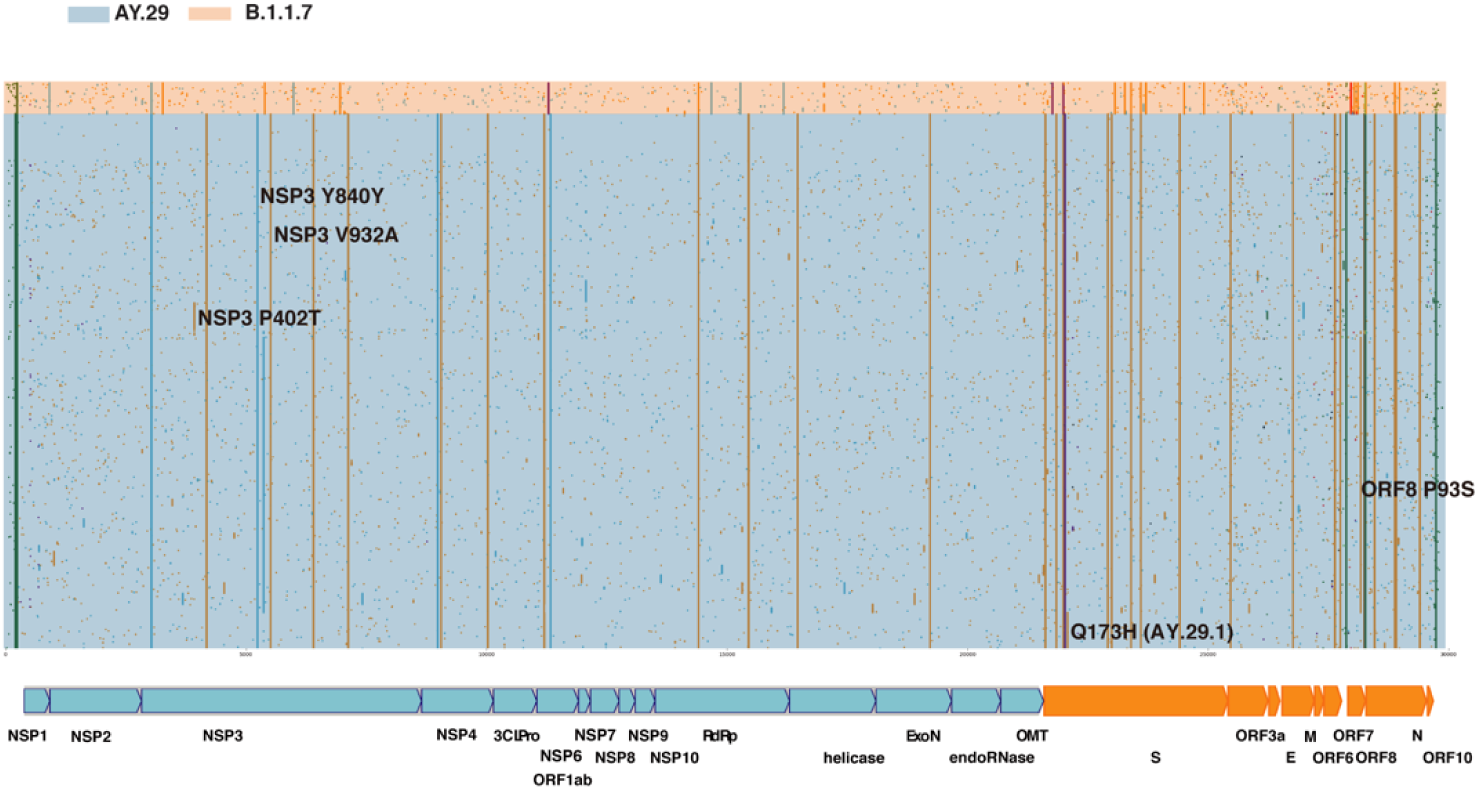
Variant Analysis of SARS-CoV-2 genomes sampled in Tokyo, Japan between July 23rd and September 5, 2021. AY.29 delta substrain is dominant in the period.

AY.29 evolved from AY.4 in Japan in April acquiring 5539C>T and 5514T>C mutations (EPI_ISL_2723567/EPI_ISL_2723568). Chronological order of emergences of 5539C>T and 5514T>C is not clear from data. The ancestral AY.4 was an exogenous strain as seen in EPI_ISL_1927416, which is captured from a traveler from India at a Japanese airport quarantine in April; however, actual introduction of the ancestral AY.4 strain might be earlier than April. In spike protein, besides D614G, L452R, T478K, P681R, and D950N, almost all AY.29 strains have T19R, T19I, G142D, and 156_158delinsG in N-terminal Domain (NTD). Among the substrains of AY.29, one with ORF8 P93S forms the largest group followed by one with spike Q173H, which is now classified as AY.29.1.

As of October 18, 2021, 98 of AY.29 exported samples are identified in 16 countries (Table 1). 44 samples in United States were found, followed by 16 samples in United Kingdom and 8 samples in Canada. From phylogenic analysis as shown in Figure 3 and ancestral strain searches, 46 distinct AY.29 strains are known to be transmitted to the outside territories of Japan. The biggest overseas AY.29 cluster, which harbors NSP3 P402T mutation, occurred in Hawaii. The introduction of the strain took place before the Olympic Games; therefore, this Hawaiian cluster is not associated with the events. Another large cluster due to AY.29 strain with ORF8 P93S and NPS3 N873D mutations seems to be related to United States Military stationed in Okinawa, the southern island prefecture; therefore, this strain is unrelated to the events as well. United States has 10 distinct AY.29 introductions, and United Kingdom has 13 of them in the second place. Other countries or regions with multiple AY.29 introductions are Germany, Canada, South Korea, and Hong Kong. The rest of the countries or regions, Italy, France, Spain, Sweden, Belgium, Peru, Australia, New Zealand, and Indonesia, have a single AY.29 strain introduced. Furthermore, there are no incidents of indirect transmissions of AY.29 strains not involving Japan.

**Table 1:**
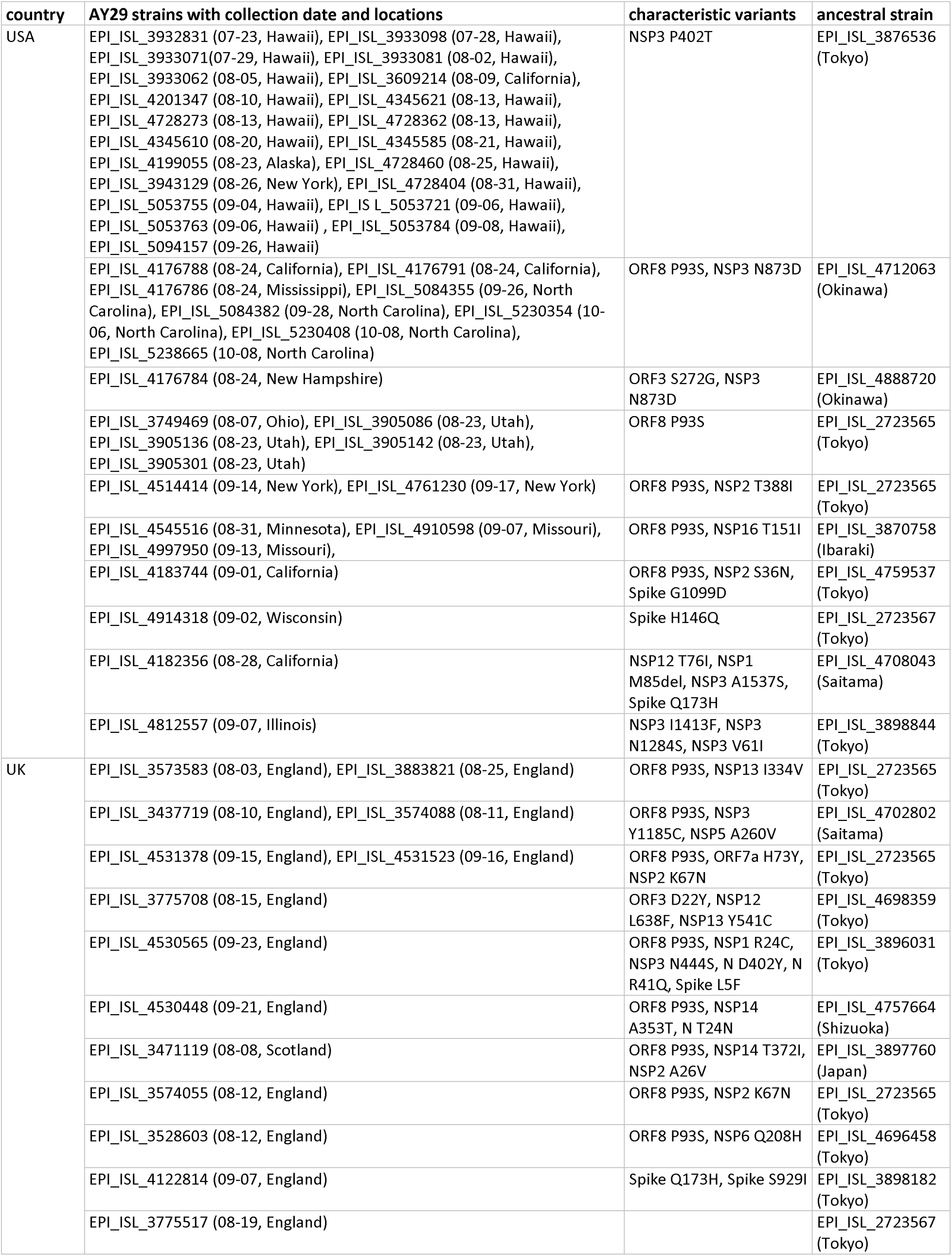
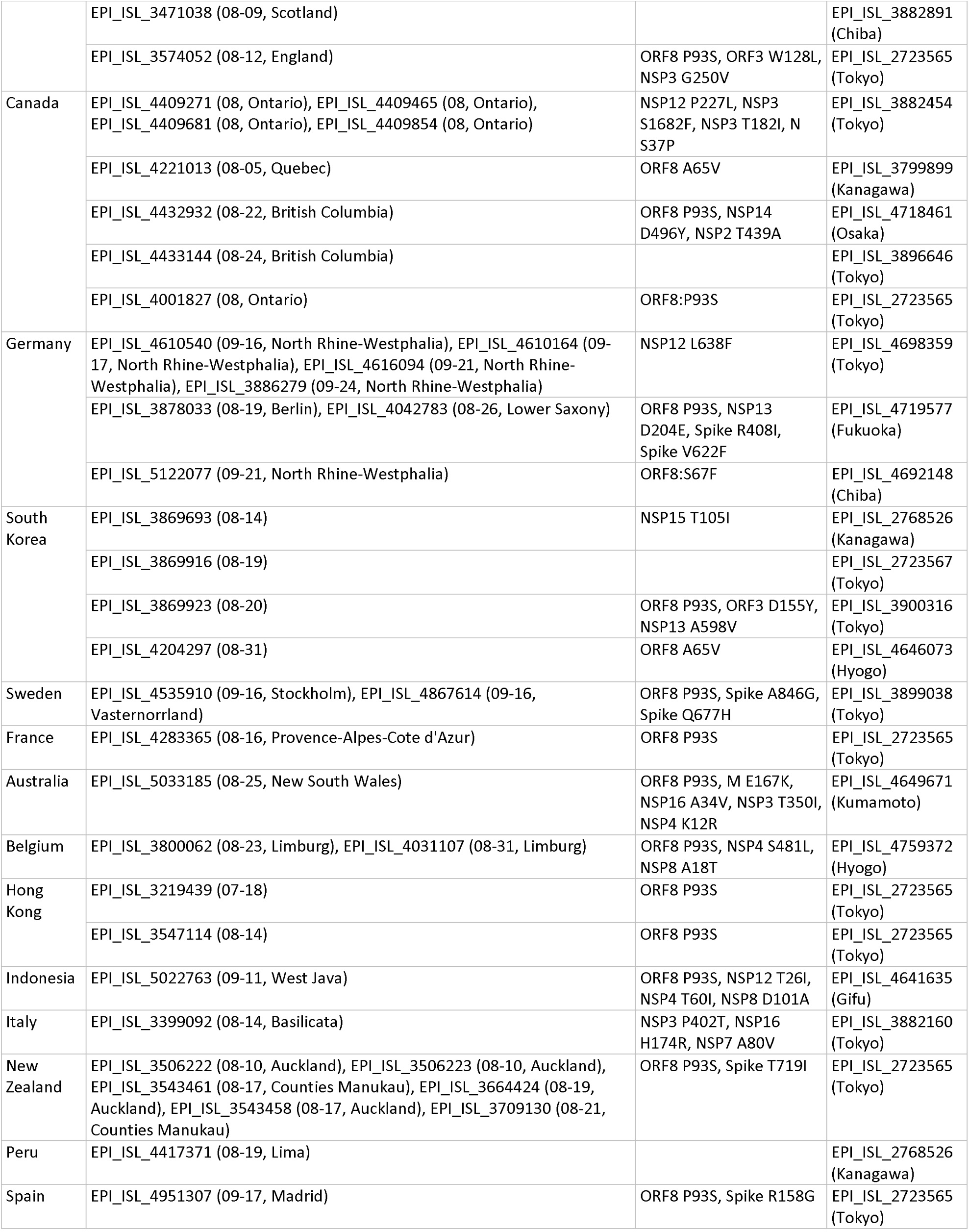
AY.29 strains identified outside of Japan. 98 samples were found in 16 countries. For each individual introduction represented by a row, additional characteristic mutations and ancestral strain in Japan are shown.

**Figure 3:**
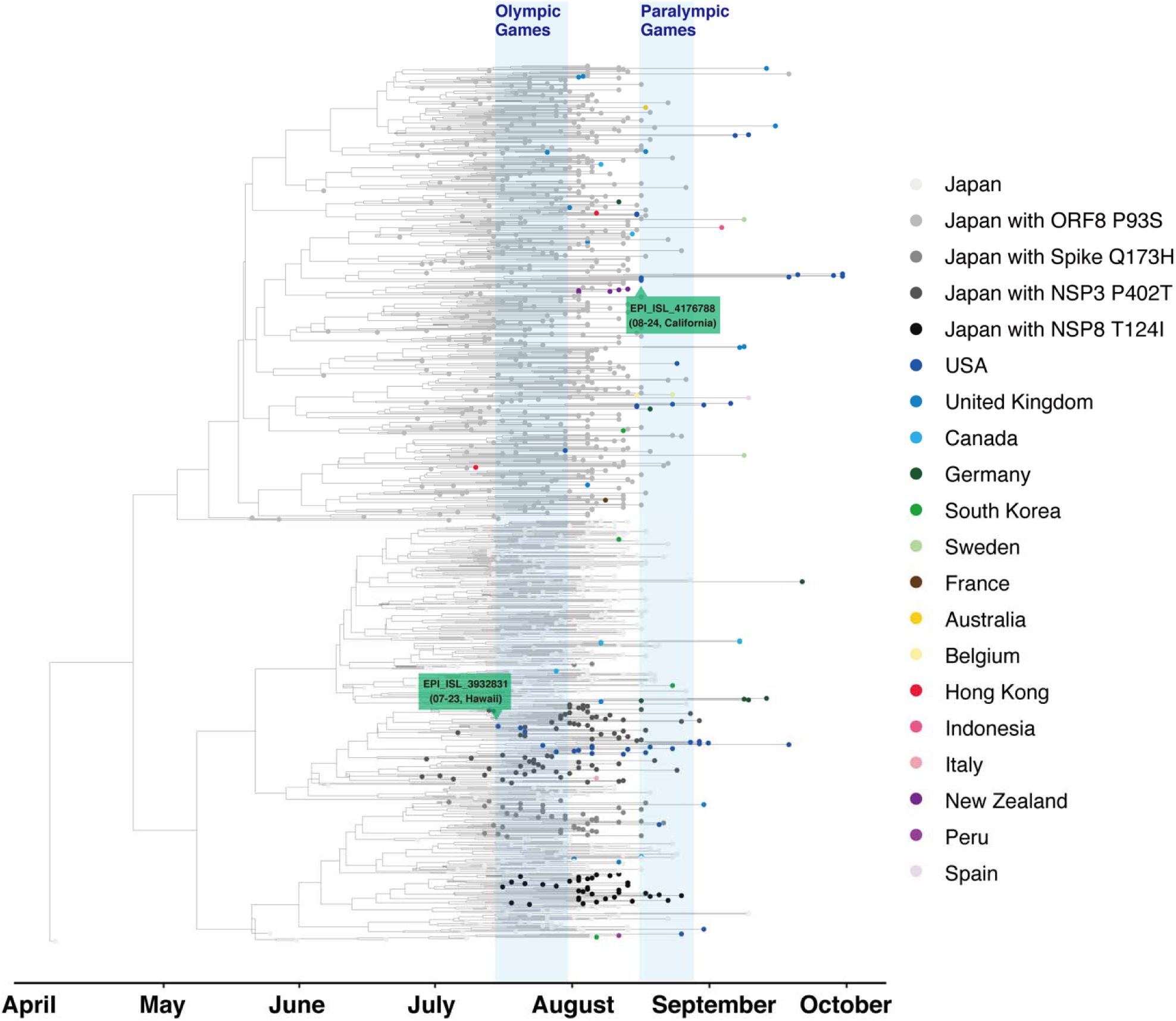
Phylogenetic analysis of AY.29 strains. All overseas samples were combined with randomly selected 1000 AY.29 genomes in Japan.

## Discussion

Although AY.29 strains have been identified outside of Japan, with limited knowledge, it is not certain how many of them were indeed associated with Olympic and Paralympic cross-border traffics. Olympic and Paralympic related travelers accounts for approximately 1/3 of entire outbound travelers from Japan as implied from Figure 1E-G; therefore, the events have likely contributed to the cross-border transmissions to some degree. To aid further investigation, it will be useful to know the actual number of the event driven cross-border transmissions. By knowing how the cross-border transmissions occurred, IOC and IPC may refine the Playbooks(19) to make the upcoming Beijing Winter Olympic Games even safer.

As vaccination rate in Japan increased during the summer, the surge caused by AY.29 started to decline. Since the number of positive cases in Japan made significant decrease from the peak in August, AY.29 strain does not impose threat to vaccinated region. However, the strain demonstrated that it is still life threatening for the unvaccinated. Although many breakthrough cases were reported due to delta variant(20, 21) and various reports that indicate compromised effectiveness against delta variant were released(22-24), it is incontrovertible that vaccines are quite effective against AY.29 strains. Therefore, leakages of AY.29 outside of Japan to vaccinated countries would not induce serious issues. Unfortunately, this might not be the case for low-income countries, where only a few percent of citizens are vaccinated against SARS-CoV-2(8). AY.29 is highly infective to unvaccinated people as demonstrated in Japan. Due to the diverse nature of summer Olympic Games with participants from 205 countries, many participants in the Tokyo Olympic Games were from countries with low vaccination rates. Particularly, many African nations have low vaccination rates. Besides, in Africa only 1 in 7 positive cases were reported according to World Health Organization (WHO)(25); therefore, it is challenging to detect any outbreaks in general. More rigorous departure screenings for participants from unvaccinated countries might have been necessary for participants from those regions. Samplings of genomes in many countries are almost none or very low according to GISAID(26). Even if they are sampled, it is common to take months before being shared. At this point, there is no sign of significant surge due to the exported AY.29 strains; however, it is important to keep monitoring of this potentially devastating strain for the time being.

## Data Availability

All genome data used in the study is listed in supplemental table and available at GISAID and NCBI. COVID-19 case and death number in japan is available at Our World in Data.

https://www.gisaid.org/

https://ourworldindata.org/

https://www.ncbi.nlm.nih.gov/

## List of Abbreviations

COVID-19: Coronavirus Disease 2019
SARS-CoV-2: Severe Acute Respiratory Syndrome Coronavirus 2
GISAID: Global initiative on sharing all influenza data
IOC: International Olympic Committee
IPC: International Paralympic Committee
VoC: Variant of Concern
VoI: Variant of Interest
WHO: World Health Organization
Harumi OV/PV: Harumi Olympic Village / Paralympic Village

## Author Contributions

T.K. conceived the idea and performed the research, analyzed the data, and wrote the manuscript. R.T. analyzed the data and wrote the manuscript. K.K. and A.Y. supported the data analysis. M.K. and S.I. supervised the study. All interpreted the data, reviewed the manuscript and made refinements.

## Acknowledgements

We thank Teruhiko Yoshida for technical assistance. We gratefully acknowledge the authors, originating and submitting laboratories of the sequences from GISAID’s EpiFlu™ Database and NCBI on which this research is based. The list of genomes from GISAID’s EpiFlu™ Database is provided in Supplemental Table 1.

## Supplemental Materials

**Supplemental Table 1: Genomes Used in Analysis**

## References

1. NHK. Tokyo Olympic and Paralympic Games Committee Reported Status of New Coronavirus (in Japanese): NHK; 2021 [Available from: https://www3.nhk.or.jp/news/html/20210929/k10013281651000.html.

2. Yomiuri Shimbun. Olympic and Paralympic Games Related Visitors were Reduced to 1/3 Excluding Athletes (in Japanese) 2021 [Available from: https://www.yomiuri.co.jp/olympic/2020/20210618-OYT1T50254.

3. Rambaut A, Holmes EC, Hill V, O’Toole Á, McCrone JT, Ruis C, et al. A dynamic nomenclature proposal for SARS-CoV-2 to assist genomic epidemiology. bioRxiv. 2020:2020.04.17.046086.

4. Michael Rajah M, Hubert M, Bishop E, Saunders N, Robinot R, Grzelak L, et al. SARS-CoV-2 Alpha, Beta and Delta variants display enhanced Spike-mediated Syncytia Formation. EMBO J. 2021:e108944.

5. Current Situation of Infection, September 1, 2021. National Institute of Infectious Disease; 2021.

6. The Tokyo Organising Committee of the Olympic and Paralympic Games. COVID-19 Positive Case List.; 2021 September 8.

7. Wells CR, Townsend JP, Pandey A, Moghadas SM, Krieger G, Singer B, et al. Optimal COVID-19 quarantine and testing strategies. Nature Communications. 2021;12(1):356.

8. Coronavirus Pandemic (COVID-19) [Internet]. OurWorldInData.org. 2020.

9. Shu Y, McCauley J. GISAID: Global initiative on sharing all influenza data - from vision to reality. Euro Surveill. 2017;22(13).

10. Elbe S, Buckland-Merrett G. Data, disease and diplomacy: GISAID’s innovative contribution to global health. Global Challenges. 2017;1(1):33–46.

11. Koyama T. PD, Parida L. Variant analysis of SARS-CoV-2 genomes. Bull World Health Organ. 2020.

12. Needleman SB, Wunsch CD. A general method applicable to the search for similarities in the amino acid sequence of two proteins. Journal of Molecular Biology. 1970;48(3):443–53.

13. Katoh K, Misawa K, Kuma K, Miyata T. MAFFT: a novel method for rapid multiple sequence alignment based on fast Fourier transform. Nucleic Acids Res. 2002;30(14):3059–66.

14. Hasegawa M, Kishino H, Yano T. Dating of the human-ape splitting by a molecular clock of mitochondrial DNA. J Mol Evol. 1985;22(2):160–74.

15. Yu G. Using ggtree to Visualize Data on Tree-Like Structures. Curr Protoc Bioinformatics. 2020;69(1):e96.

16. Tokumasu R, Weeraratne D, Snowdon J, Parida L, Kudo M, Koyama T. Introductions and evolutions of SARS-CoV-2 strains in Japan. medRxiv. 2021:2021.02.26.21252555.

17. Sanderson T, Barrett JC. Variation at Spike position 142 in SARS-CoV-2 Delta genomes is a technical artifact caused by dropout of a sequencing amplicon. medRxiv. 2021:2021.10.14.21264847.

18. Abe T, Arita M. Genomic Surveillance in Japan of AY. 29—A New Sub-lineage of SARS-CoV-2 Delta Variant with C5239T and T5514C Mutations. medRxiv. 2021:2021.09.20.21263869.

19. International Olympic Committee. Tokyo 2020 Playbooks 2021 [Available from: https://olympics.com/ioc/tokyo-2020-playbooks.

20. Brown CM, Vostok J, Johnson H, Burns M, Gharpure R, Sami S, et al. Outbreak of SARS-CoV-2 Infections, Including COVID-19 Vaccine Breakthrough Infections, Associated with Large Public Gatherings - Barnstable County, Massachusetts, July 2021. MMWR Morb Mortal Wkly Rep. 2021;70(31):1059-62. 21.

21. Herlihy R, Bamberg W, Burakoff A, Alden N, Severson R, Bush E, et al. Rapid Increase in Circulation of the SARS-CoV-2 B.1.617.2 (Delta) Variant - Mesa County, Colorado, April-June 2021. MMWR Morb Mortal Wkly Rep. 2021;70(32):1084–7.

22. Planas D, Veyer D, Baidaliuk A, Staropoli I, Guivel-Benhassine F, Rajah MM, et al. Reduced sensitivity of SARS-CoV-2 variant Delta to antibody neutralization. Nature. 2021;596(7871):276–80.

23. Fowlkes A, Gaglani M, Groover K, Thiese MS, Tyner H, Ellingson K, et al. Effectiveness of COVID-19 Vaccines in Preventing SARS-CoV-2 Infection Among Frontline Workers Before and During B.1.617.2 (Delta) Variant Predominance - Eight U.S. Locations, December 2020-August 2021. MMWR Morb Mortal Wkly Rep. 2021;70(34):1167–9.

24. Mlcochova P, Kemp SA, Dhar MS, Papa G, Meng B, Ferreira IATM, et al. SARS-CoV-2 B.1.617.2 Delta variant replication and immune evasion. Nature. 2021.

25. World Health Organization. Six in seven COVID-19 infections go undetected in Africa 2021 [Available from: https://www.afro.who.int/news/six-seven-covid-19-infections-go-undetected-africa.

26. GISAID. 2021 [Available from: https://www.gisaid.org/index.php?id=208.

